# Automated CTA-Based Assessment of Aortic Remodeling Through Volumetric Biomarkers in Type B Aortic Dissection

**DOI:** 10.64898/2026.11.23.26368738

**Authors:** Alice Maccarini, Marco Ferraresi, Alessandro Molinari, Janko Jovanovic, Giovanni Rossi, Luca Mainardi, Pietro Cerveri

**Affiliations:** Università di Pavia, Via A. Ferrata, 5, Pavia, 27100, Italy; Department of Electronics, Information and Bioengineering, Politecnico di Milano University, via Ponzio 34/5, Milan, 20133, Italy; Cardiothoracic and Vascular Department, Unit of Vascular Surgery, Fondazione IRCCS Policlinico San Matteo, Viale Camillo Golgi, 19, Pavia, 27100, Italy

**Author notes:** Corresponding author at: University of Pavia, Pavia 27100, Italy Email address (Pietro Cerveri).

**Keywords:** Type B aortic dissection, Deep learning, Computed Tomographic Angiography, TEVAR, Volumetric analysis

## Abstract

Type B aortic dissection (TBAD) requires lifelong morphological monitoring using computed tomography angiography (CTA). TBAD separates blood flow into the true lumen (TL), the original aortic channel, and the false lumen (FL), created by the dissection. Although three-dimensional volumetric analysis may complement diameter-based surveillance, its clinical adoption is limited by the time required for manual segmentation, particularly in postoperative scans affected by metallic artifacts. This study developed and evaluated an automated CTA-based framework for volumetric surveillance of TBAD across native and post-TEVAR conditions. We propose AORTASCAN, a hierarchical multi-stage framework for automated segmentation of the aorta, TL, and FL, extended to stented anatomy through a metal-aware triple-input strategy. The system computes conventional diameter measurements and volumetric biomarkers, including the perfusion dissection index (PDI), defined as the ratio between perfused FL volume and total perfused aortic volume. Domain adaptation was achieved using transfer learning and partial-label learning across heterogeneous clinical annotations. In native cases, AORTA-SCAN achieved a global Dice score of 0.84 and significantly improved FL segmentation compared with the nnU-Net baseline (0.84 vs 0.71, *p <* 0.001). Volumetric analysis showed high agreement with expert references, with a mean PDI bias of *−*0.17%. In postoperative cases, TL segmentation remained robust (Dice = 0.90), while FL and stent Dice were 0.75 and 0.72, respectively. Processing was completed in less than 1 minute per case on a standard workstation. AORTA-SCAN enables automated volumetric surveillance of TBAD and supports scalable extraction of PDI and diameter biomarkers from native and post-TEVAR CTA.

## 1. Introduction

Type B aortic dissection (TBAD) is a life-threatening cardiovascular condition routinely evaluated and monitored using computed tomography angiography (CTA), which represents the clinical standard imaging modality for diagnostic assessment ([9]). TBAD is characterised by an intimal tear that allows blood to enter the aortic wall, creating a false lumen (FL) alongside the true lumen (TL) ([10]). In uncomplicated cases, management is predominantly conservative and relies on lifelong imaging surveillance ([1]), whereas thoracic endovascular aortic repair (TEVAR) or the STABILISE technique are indicated in the presence of complications such as malperfusion or impending rupture, and may also be considered in selected high-risk patients [20, 8]. Current clinical decision-making and evaluation of aortic remodeling are largely based on the extent of the dissection, false lumen patency, total aortic diameter, and dimensional changes in both lumina ([19]). Although these metrics are clinically established, they provide only a partial representation of the disease, failing to capture the full spatial complexity of aortic remodeling and offering limited predictive value for late aortic events. The conventional definition of “complete remodeling” is dichotomous and based on complete false lumen thrombosis or disappearance, whereas “partial” or “positive remodeling” encompasses a wide spectrum of heterogeneous morphological changes ([10]). To better characterise these intermediate conditions, increasing attention has been directed toward volumetric and geometric descriptors of aortic dissection.

Several studies have demonstrated the potential of volumetric analysis. Sobocinski et al. quantified true and false lumen volumes to compare surgical strategies ([25]), while subsequent works showed that volumetric parameters can predict long-term aortic events ([5, 13]). In particular, the perfusion dissection index (PDI), defined as the ratio between perfused FL volume and total perfused aortic volume, has been suggested as a robust biomarker of aortic remodeling and a predictor of reintervention ([11]). Despite this growing evidence, the clinical adoption of volumetric analysis remains limited by the need for accurate segmentation of aortic lumen compartments, a process that is time-consuming when performed manually, typically requiring 60–75 minutes per scan. This limitation is further exacerbated in postoperative scenarios, where stent-induced artifacts and altered vascular geometry complicate image interpretation and delay quantitative assessment ([24]). Deep learning has enabled substantial progress in automated image processing ([23]), and especially in aortic segmentation, with approaches achieving high accuracy in delineating TL and FL on preoperative CTA datasets ([3, 7, 14, 18, 28]). These methods, based on convolutional neural networks and more recently, transformer-based architectures, enable fast and accurate morphological analysis. However, segmentation accuracy alone does not necessarily guarantee reliable extraction of clinically relevant volumetric biomarkers. Overall, a significant translational gap persists. Most existing methods treat TBAD analysis primarily as a conventional segmentation problem, optimizing voxel-wise overlap metrics on curated datasets. However, longitudinal clinical surveillance requires substantially more than accurate contour delineation. Robust deployment in real-world settings demands anatomical consistency across heterogeneous domains, preservation of volumetric biomarker fidelity, and resilience to postoperative alterations such as stent-induced beam-hardening artifacts. These challenges remain largely unaddressed by current monolithic segmentation pipelines. As a result, no unified framework enables robust, clinically deployable analysis of TBAD across both native and post-TEVAR settings. In this work, we propose AORTA-SCAN, a hierarchical and domain-adaptive framework for volumetric surveillance of primary and residual TBAD. Unlike conventional end-to-end segmentation pipelines, the proposed approach explicitly decouples anatomical localization from pathological refinement, enabling robust segmentation of the TL and FL together with volumetric analysis across heterogeneous CTA domains, including post-TEVAR anatomy. The main contributions of this study are threefold. First, we introduce a hierarchical segmentation strategy that separates global anatomical localization of the aorta from fine-grained lumen analysis through a probabilistic soft-prior, improving robustness and computational efficiency. Second, we implement a domain adaptation scheme based on transfer learning and partial-label learning to mitigate distribution shifts between curated datasets and heterogeneous real-world clinical data. Third, we extend the framework to postoperative scenarios by incorporating a metal-aware input representation and explicitly modeling the stent as a topological anatomical constraint, enabling reliable partitioning of intra-stent and extra-stent volumes despite severe beam-hardening artifacts. Importantly, the proposed framework is designed not only to optimize voxel-wise segmentation accuracy but also to preserve the stability and reproducibility of clinically relevant volumetric biomarkers across heterogeneous clinical domains. By integrating these components into a unified pipeline, AORTA-SCAN enables automated extraction of volumetric descriptors, including TL and FL volumes and the PDI, directly from routine CTA scans.

## 2. Related Works

Early work by Cao et al. demonstrated the feasibility of fully automated TBAD segmentation in CTA using convolutional neural networks ([3]), while subsequent multi-stage frameworks have further improved segmentation performance and enabled volumetric analysis ([7]). More recently, UNet-based architectures have achieved high accuracy in segmenting aortic dissection components; however, these studies have generally been limited by relatively small datasets and constrained generalizability ([14]). Integrated deep learning pipelines have also been described to jointly perform aortic segmentation, dissection detection, TL/FL segmentation, and Stanford classification, achieving high accuracy across multiple tasks ([28]). While these approaches demonstrate the potential of end-to-end automation for comprehensive analysis, they remain primarily focused on classification and segmentation performance, without addressing longitudinal volumetric quantification and biomarker extraction. In parallel, deep-learning-based segmentation methods have enabled automated quantification of aortic diameter and dissection components, including TL, circulating FL, and thrombus ([12]). Notably, local FL volumes measured near the maximal aortic diameter have been shown to outperform global volumetric measures and diameter-based metrics in predicting adverse aortic remodeling, emphasizing the importance of spatially resolved volumetric descriptors. More broadly, artificial intelligence-assisted approaches have demonstrated strong performance in the automated detection of acute aortic syndromes (AAS) across large multicenter datasets, supporting their potential role in rapid clinical triage ([6]). Complementary strategies have also explored the automatic detection of anatomical landmarks for longitudinal assessment. In particular, deep reinforcement learning (DRL) approaches have demonstrated high accuracy in localizing aortic landmarks in TBAD ([2]). However, these methods are inherently limited to sparse geometric representations and do not capture the full volumetric distribution of luminal compartments required for comprehensive morphological and volumetric characterization. Collectively, these findings highlight the need for robust frameworks capable of integrating accurate segmentation, volumetric biomarker extraction, and postoperative robustness across heterogeneous clinical domains.

## 3. Methods

### 3.1. Study Design and Data Sources

The framework was developed through four sequential stages using five arterial-phase CTA datasets with complementary anatomical coverage and annotation characteristics (Table 1, Figure 1). This staged design was adopted to separate coarse anatomical localization from fine luminal segmentation and to progressively adapt the model from curated public datasets to heterogeneous real-world clinical data. Such a strategy was particularly important because the number of locally annotated clinical CTA scans was limited, especially in the postoperative setting, making standalone training on the ASST Lecco cohort insufficiently robust. In SEG1, the AortaSeg24 ([16]) dataset was used to train a binary aortic localization model based on a U-Net-like architecture capable of identifying the thoraco-abdominal aortic tract and defining a subject-specific region of interest for downstream analysis. In SEG2, the ImageTBAD dataset ([26]) was used to train an initial TL/FL segmentation model on the localized aortic subvolume. A native lumen refinement stage was subsequently performed in SEG3. First, the model was refined using the Graz dataset ([22]), which provided broader thoracoabdominal anatomical coverage than ImageTBAD. Second, preoperative domain adaptation was performed by continuing refinement using the Graz dataset together with the ASST Lecco preoperative cohort. This combined refinement strategy preserved the broader anatomical coverage available in the Graz annotations, including luminal extensions into branch vessels, while simultaneously adapting the model to the heterogeneous imaging characteristics of the ASST Lecco cohort. The final postoperative stage, SEG4, extended the framework to stented anatomy using the ASST Lecco postoperative cohort, composed of 21 cases. Due to inadequate contrast enhancement for arterial lumen delineation and volumetric surveillance, one venousphase examination was excluded. The final SEG4 dataset thus comprised 20 arterial-phase CTA scans, establishing arterial-phase acquisition as a mandatory baseline requirement for the reliable execution of the AORTA-SCAN framework. Among these, 9 patients had paired preoperative CTA scans available for longitudinal analysis. SEG2, together with its refinement stages SEG3 and SEG4, was implemented using a custom attention-enhanced residual 3D U-Net backbone described in Section 3.4. ASST Lecco data, provided in pseudo-anonymized form, were acquired under the Ethical committed approval entitled STABILISE REGISTRY n. 529, 2019 (Regione Lombardia ASST Lecco). The postoperative ASST Lecco dataset is publicly available at https://drive.google.com/drive/folders/1pGjqeXYNm_SI0jprhsuw0Wre97ARkgrR?usp=sharing.

**Figure 1:**
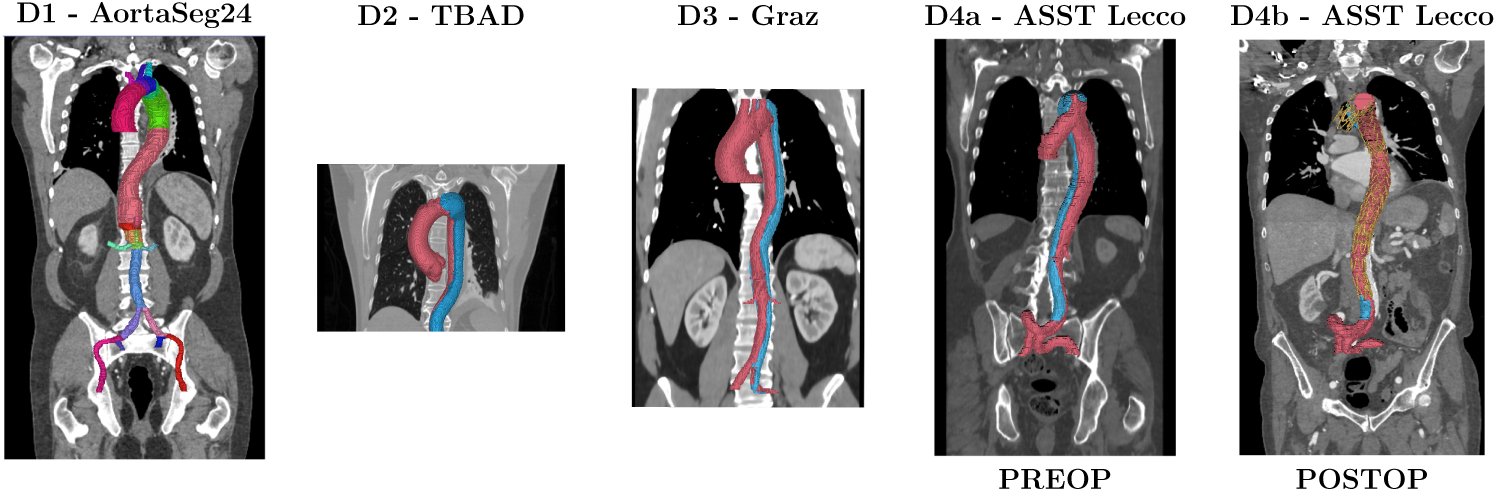
Five available datasets. From left to right: AortaSeg24, ImageTBAD, Graz University, preoperative ASST Lecco, postoperative ASST Lecco.

**Table 1:** Characteristics of datasets used for model development, including annotation type and anatomical coverage. TL: true lumen, FL: false lumen, B: background, TH: thoracic, AB: abdominal.

| **Dataset** | **N** | **Setting** | **Annotations** | **Anatomical Region** | **Stage** |
|---|---|---|---|---|---|
| D1: AortaSeg24 ([16]) | 50 | PRE | Aorta | TH + AB | SEG1 |
| D2: ImageTBAD ([26]) | 100 | PRE | TL, FL, B | TH | SEG2 |
| D3: Graz ([22]) | 40 | PRE | TL, FL, B | TH + AB | SEG3 |
| D4a: ASST Lecco | 10 | PRE | TL, FL, B | TH + AB | SEG3 |
| D4b: ASST Lecco | 20 | POST | TL, FL, B, S | TH + AB | SEG4 |

**Table 2:** Role of each dataset in the multi-stage framework, including training, refinement, and evaluation phases.

| **Dataset** | **Stage** | **Role** | **Usage** |
|---|---|---|---|
| D1 | SEG1 | Aorta localization | Training |
| D2 | SEG2 | TL/FL coarse (PRE) | Training |
| D3 + D4a | SEG3 | TL/FL refinement (PRE) | Fine-tuning |
| D4b | SEG4 | TL/FL/Stent (POST) | Fine-tuning |

### 3.2. Domain Adaptation Strategy

The integration of heterogeneous arterial-phase CTA datasets required a unified annotation and domain adaptation strategy to ensure morphological consistency across native and stented domains. In the AortaSeg24 dataset, the original multi-class annotations were consolidated into a single aortic mask to support robust anatomical anchoring during coarse localization. In the ImageTBAD dataset, false-lumen thrombus annotations, when present, were merged into the FL class to preserve volumetric consistency during TL/FL quantification. The ImageTBAD and Graz datasets provided voxel-wise annotations of TL, FL, and background, which were used as reference standards for supervised training and refinement. In contrast, the local ASST Lecco annotations were generated within a clinical workflow and were restricted to the aortic channel, without the extended branch-vessel coverage available in the Graz annotations. To mitigate inter-observer variability and improve adaptation to heterogeneous clinical annotations, a transfer learning and partial-label learning strategy was adopted. Lecco annotations were therefore treated as partial labels during refinement, allowing the model to adapt to the local imaging domain while avoiding penalization of anatomically present but unlabeled regions. For postoperative scans, explicit stent annotations were unavailable. Therefore, metallic regions were identified using an anatomically constrained threshold-based strategy. The coarse aortic mask was first used to restrict the candidate search region, after which metallic voxels were identified using a high-intensity threshold (*>* 600 HU), exploiting the characteristic attenuation of endovascular devices in CTA. Despite blooming artifacts and partial-volume effects, this approach provided a reproducible anatomical reference for stent localization and postoperative lumen partitioning across heterogeneous postoperative datasets.

### 3.3. Data Preprocessing

To ensure consistency across datasets with heterogeneous acquisition protocols and annotation characteristics, all CTA volumes underwent a standardized preprocessing pipeline implemented using the MONAI framework ([4]). Volumes were resampled to an isotropic resolution of 1.0 *×* 1.0 *×* 1.0 mm using bilinear interpolation and reoriented to a standard Right-AnteriorSuperior (RAS) coordinate system. Preprocessing strategies were adapted according to the specific requirements of the hierarchical framework and the imaging domain. For the initial coarse aortic localization stage, image intensities were clipped to a broad vascular range (*−*200 to +600 HU) to enhance contrast between the aorta and surrounding soft tissues. For fine-grained lumen segmentation, a domain-specific multi-windowing approach was implemented to maximize feature extraction. In preoperative scans, a dual-channel input representation was adopted, consisting of a vascular intensity channel, clipped to *−*200 to +1500 HU and normalized using 0.5%–99.5% percentile scaling with adaptive histogram equalization, together with a probabilistic Gaussian soft-prior derived from the coarse aortic segmentation. In postoperative scans, this representation was extended to a triplechannel input by including a metal-sensitive channel. This third channel was obtained using high-intensity thresholding (+600 to +2000 HU), enabling explicit localization of metallic structures and mitigating beam-hardening artifacts. Furthermore, given the limited number of postoperative cases and the sparse spatial distribution of metallic stent voxels, the additional metal-sensitive channel was introduced to improve representation of metallic structures during training. All input channels were subsequently normalized using instance-wise Z-score normalization to ensure consistent feature scaling across different CT scanners.

### 3.4. Hierarchical Geometry-Aware Framework

#### 3.4.1. Model Architecture

The AORTA-SCAN framework was trained as a hierarchical multi-stage segmentation process designed to address the main challenges of TBAD imaging, including anatomical localization, heterogeneous luminal morphology, and postoperative metallic artifacts (Fig. 2). During training, the framework followed a spatial cascade in which coarse global anatomical information progressively guided fine pathological refinement. In the first stage, a coarse anatomical localization module based on a U-Net-like architecture was trained to identify the thoraco-abdominal aortic tract and define a subjectspecific region of interest. The coarse target mask was defined as the union of all available aortic anatomical labels and dissection-related structures, providing a robust anatomical envelope for downstream localization. By isolating the aorta from surrounding anatomical structures, this stage reduced background variability and constrained the downstream analysis to the relevant vascular anatomy. A probabilistic Gaussian soft-prior derived from the coarse aortic segmentation was subsequently propagated to the downstream high-resolution modules as an additional anatomical guidance channel. The prior was defined as a spatial Gaussian distribution centered on the centroid of the coarse aortic mask:

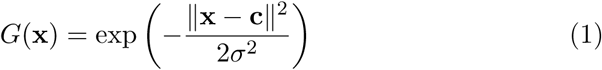

**Figure 2:**
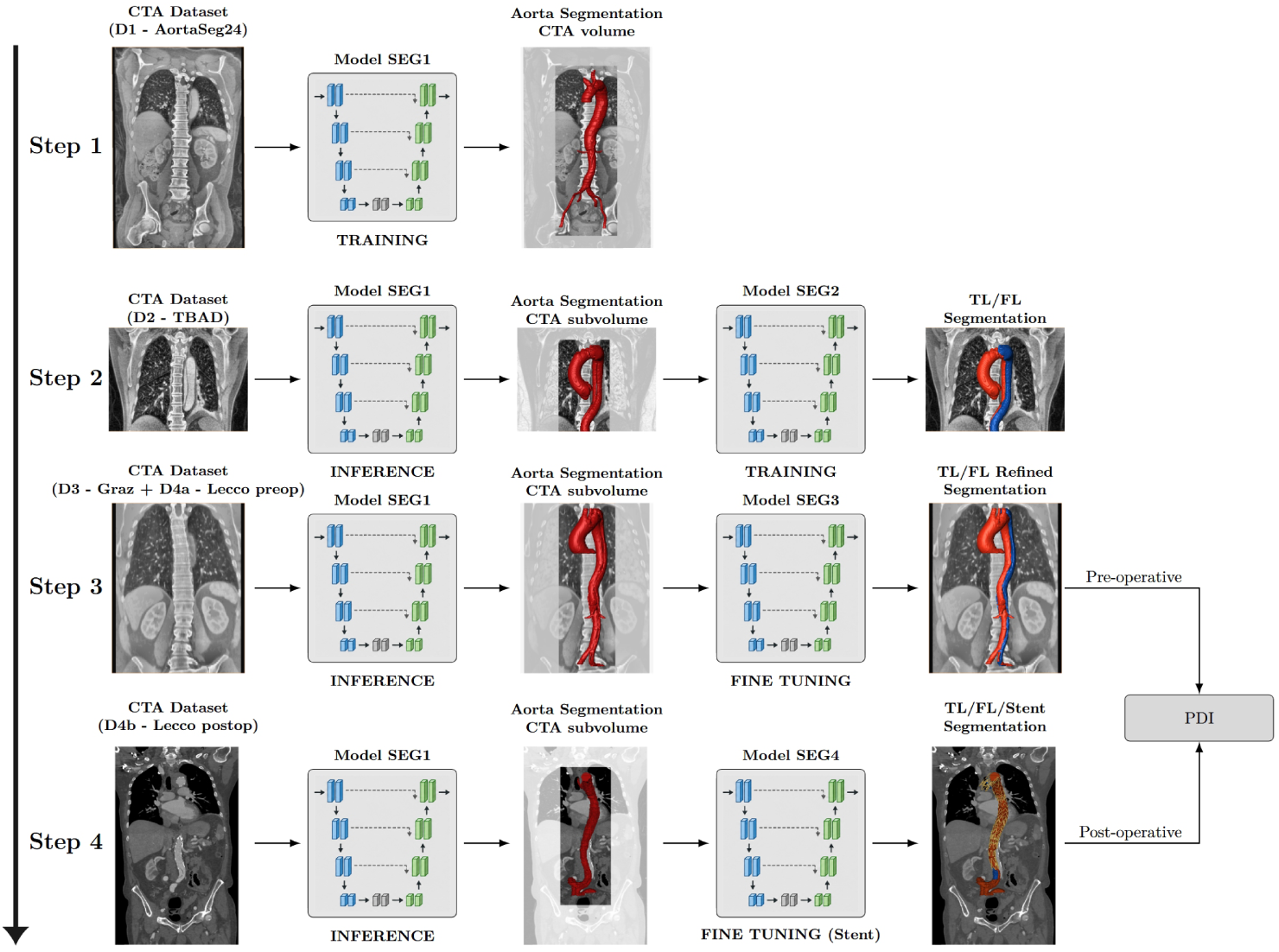
Overview of the hierarchical AORTA-SCAN pipeline for automated volumetric surveillance of native and post-TEVAR TBAD. The framework progressively combines coarse anatomical localization, fine-grained lumen refinement, domain adaptation, and stent-aware segmentation for volumetric biomarker extraction.

where **x** denotes the voxel coordinate, **c** the centroid of the coarse aortic segmentation, and *σ* controls the spatial spread of the prior. In this study, *σ* was empirically fixed to 0.3 relative to the normalized dimensions of the localized aortic subvolume. Testing values within the 0.2–0.4 range yielded no substantial differences in segmentation performance. By providing a smooth spatial weighting centered on the aortic lumen, the soft-prior reduced background ambiguity and supported more stable delineation of TL and FL boundaries during training. The fine-grained segmentation module used in SEG2, SEG3, and SEG4 was implemented using a custom attentionenhanced residual 3D U-Net backbone (Fig. 3). The network followed a symmetric encoder–decoder architecture with skip connections between corresponding resolution levels. Each encoder stage consisted of two residual convolutional blocks followed by an attention-guided downsampling operation. Feature channels were progressively increased with depth, starting from an initial number of filters and doubling at each resolution level. At each skip connection, squeeze-and-excitation (SE) blocks recalibrated channelwise feature responses before fusion with the decoder pathway. The decoder performed progressive upsampling using interpolation followed by convolution, concatenated the corresponding attention-refined encoder features, and applied residual convolutional refinement. A final 1 *×* 1 *×* 1 convolution mapped the decoded representation to the target segmentation classes. Group normalization was adopted within convolutional blocks to improve training stability for patch-based 3D inputs and limited batch sizes. The same backbone architecture was used across native and postoperative configurations, with the input dimensionality and output label space adapted to the imaging domain. In native TBAD scans, the network received a dual-channel input composed of the vascular CTA intensity image and the Gaussian anatomical soft-prior generated by the coarse localization stage, producing TL and FL predictions. In postoperative scans, the framework was extended to a triple-channel configuration by incorporating an additional metal-sensitive intensity channel (*>* 600 HU), enabling explicit exploitation of stent-related information while preserving the same encoder– decoder structure. In this configuration, the output layer was adapted to predict TL, FL, and stent classes. The explicit stent representation acted as a topological constraint for postoperative lumen partitioning, reducing ambiguity caused by metallic blooming and beam-hardening artifacts during volumetric analysis.

**Figure 3:**
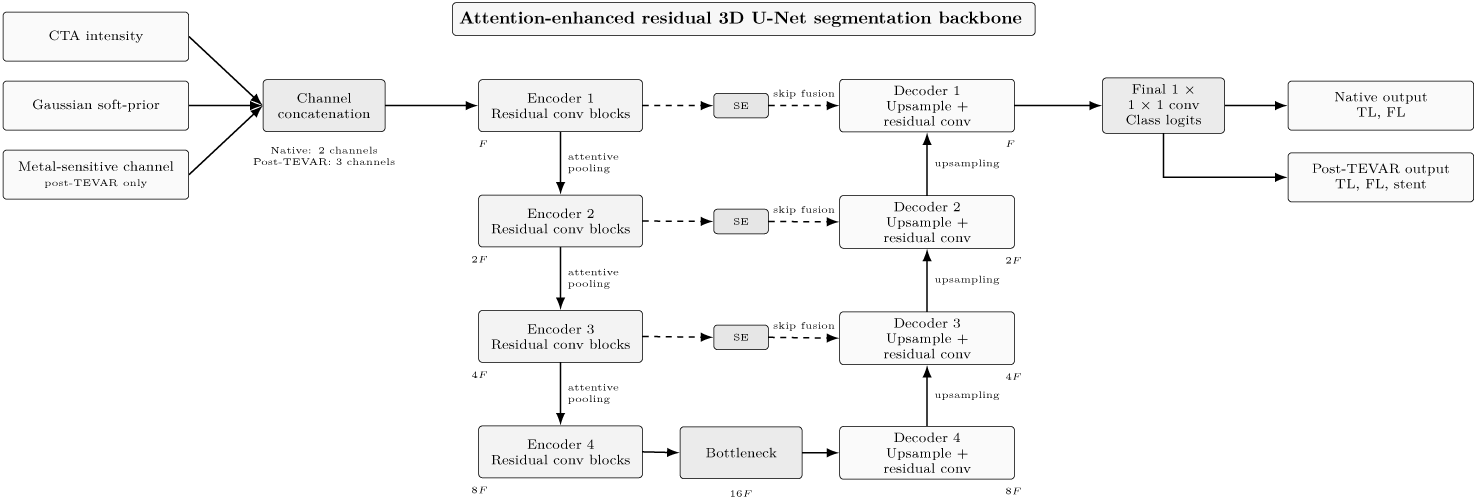
Architecture of the proposed attention-enhanced residual 3D U-Net backbone used for fine-grained TL/FL segmentation. Native and postoperative configurations differed in the number of input channels and output classes. SE blocks indicate squeeze-and-excitation channel recalibration modules, while *F* denotes the base number of feature channels progressively doubled across encoder levels.

#### 3.4.2. Training Strategy and Implementation Details

To improve generalization and reduce overfitting, on-the-fly 3D data augmentation was applied during training. This included random extraction of 128^3^ voxel patches, spatial transformations (axis flipping and 90*^◦^* rotations, both with *p* = 0.4), and intensity augmentations including random contrast adjustment (*γ ∈* [0.5, 1.5]), Gaussian noise (*σ* = 0.1), and Gaussian smoothing (*σ ∈* [0.5, 1.0]). Intensity-based augmentations were applied exclusively to image channels to preserve the structural integrity of the softprior input. To address class imbalance between lumen structures, stent voxels, and background, training was performed using a patch-based sampling strategy with a fixed input size of 128 *×* 128 *×* 128 voxels. Patches were extracted using MONAI label-guided cropping, which increased the probability of sampling regions containing annotated foreground classes rather than uniformly sampling the full CTA volume. For native scans, foreground sampling targeted TL and FL voxels, whereas for postoperative scans, it additionally targeted stent voxels to compensate for the sparse spatial distribution of metallic structures. This strategy increased the representation of clinically relevant structures during training while preserving sufficient background context within each patch. The patch-based balancing strategy followed principles previously adopted for multi-task encoder–decoder segmentation ([21]). The AORTA-SCAN framework was implemented using the PyTorch library and the MONAI framework ([4]). All model training procedures were conducted on a workstation equipped with a single NVIDIA H100 PCIe GPU (80 GB VRAM). We used the AdamW optimizer with an initial learning rate of 1 *×* 10*^−^*^4^ and a weight decay of 1 *×* 10*^−^*^5^. The learning rate was adjusted using a cosine annealing schedule. Training was performed with a batch size of 4 and continued until early stopping based on the validation macro Dice score; therefore, the number of epochs varied across folds and training stages, with the maximum number of epochs set to 250. The choice of objective function was tailored to each stage of the hierarchical pipeline: Generalized Dice Loss was employed for the initial training and fine-segmentation stages to address class imbalance. For domain adaptation and refinement phases involving heterogeneous clinical annotations, Partial Label Dice–Cross Entropy Loss was adopted. This masked objective computes gradients exclusively for classes explicitly present in the reference standard, preventing the penalization of unannotated structures and enabling the model to learn effectively from partially labeled clinical datasets. Final model selection within each fold was determined by the highest validation macro Dice score.

#### 3.4.3. Inference Strategy

During inference, the framework operated sequentially according to the hierarchical architecture described above. First, the coarse localization model (SEG1) identified the thoraco-abdominal aortic tract and generated a binary aortic mask. This mask was used to define a subject-specific region of interest and to compute the Gaussian anatomical soft-prior propagated to the downstream segmentation stages. The localized subvolume, together with the anatomical soft-prior, was subsequently processed by the lumen segmentation network. In native CTA scans, the inference pipeline used the dualchannel configuration to generate TL and FL predictions. In postoperative CTA scans, the additional metal-sensitive channel was included to support stent-aware segmentation and postoperative lumen partitioning. The final output consisted of voxel-wise TL, FL, and, when applicable, stent segmentations, which were subsequently used for volumetric and geometric biomarker extraction.

### 3.5. Volumetric Analysis

To translate voxel-wise segmentations into standardized clinical metrics, we developed a graph-based topological analysis module designed to provide objective morphometric data. Initially, the 3D lumen centerline was extracted via skeletonization of the predicted binary aortic mask. The absolute volumes of the TL (*V_T L_*) and FL (*V_F L_*) were then computed for comparison against the manual reference standard. To ensure a geometrically stable assessment of vessel caliber that remains robust to aortic tortuosity, we calculated the Equivalent Diameter (*D_eq_*) for each 1 mm segment along the extracted centerline. This metric is derived from the cross-sectional area (*A*) of the aortic mask perpendicular to the centerline according to the following equation:

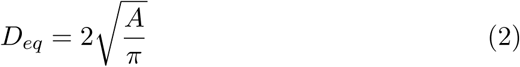

Finally, we calculated the PDI to quantify the relative disease burden and luminal compression. The PDI is defined as the ratio of the FL volume to the total aortic volume (*V_total_* = *V_T L_* + *V_F L_*), providing a 3D digital biomarker for hemodynamic risk:

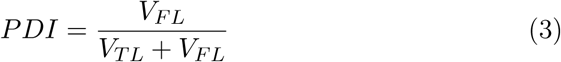

The index was computed both globally, over the entire segmented aortic volume, and regionally within predefined anatomical zones, namely Thoracic, Visceral, and Aorto-iliac, to capture spatial variations in FL dominance along the aorta. When paired preoperative and postoperative scans were available for the same patient, TL and FL volumes were additionally compared longitudinally to quantify postoperative volumetric remodeling.

To assess clinical concordance beyond voxel-wise overlap, the relative volumetric error (RVE) was computed for each predicted lumen as follows:

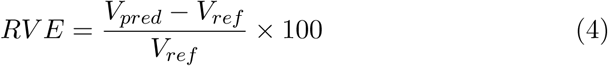

where *V_pred_*represents the predicted volume and *V_ref_* the volume of the reference standard. Unlike traditional overlap metrics such as the Dice score, RVE provides a direct assessment of the framework’s ability to preserve the physical scale of the aortic compartments, which is essential for longitudinal clinical monitoring.

### 3.6. Evaluation Metrics

Model performance was evaluated using a multi-level validation strategy assessing both voxel-wise segmentation accuracy and the clinical reliability of derived volumetric biomarkers. Segmentation performance was quantified using the Dice coefficient, Intersection over Union (IoU), sensitivity, precision, and specificity, computed at the patient level. Clinical agreement was assessed through linear regression and Bland–Altman analysis of true lumen volume (*V_T L_*), false lumen volume (*V_F L_*), and Perfusion Dissection Index (PDI). In addition, the Relative Volumetric Error (RVE) was calculated to quantify proportional discrepancies between predicted and reference volumes. Robustness was evaluated across heterogeneous datasets with different acquisition protocols and annotation schemes, including public benchmarks and real-world clinical cohorts. For partially annotated clinical datasets, partial-label learning was adopted during training, while masked evaluation metrics were used to restrict performance assessment to explicitly annotated regions. Validation was performed using a 5-fold cross-validation scheme with strict patient-level separation (80% training, 20% testing per fold) to prevent data leakage. Final results are reported as the mean across folds. Continuous variables were reported as means *±* standard deviation. Statistical comparisons between AORTA-SCAN and the baseline model were performed using the paired Wilcoxon signed-rank test. Differences were considered statistically significant for *p <* 0.05.

## 4. Experimental Results

### 4.1. Preoperative cohorts

#### 4.1.1. Segmentation Results

The segmentation framework was evaluated against a conventional nnUNet ([17]) baseline operating without hierarchical localization or anatomical soft-prior guidance (Table 3). The resulting out-of-fold predictions from the five-fold cross-validation achieved strong voxel-level segmentation performance for both luminal compartments. The Dice score improved from 0.754 to 0.816 for the TL (*p* = 3.01 *×* 10*^−^*^5^) and from 0.707 to 0.837 for the FL (*p* = 5.26 *×* 10*^−^*^8^). Similar performance increases were observed for IoU, precision, and recall across both classes. For the TL, precision and recall increased from 0.810 and 0.732 in the baseline to 0.853 and 0.796, respectively. The largest improvement was observed in the FL, where precision increased from 0.722 to 0.819 and recall from 0.720 to 0.875. Overall, these results indicate that the hierarchical refinement strategy improved both boundary reliability and lumen completeness relative to the baseline. In particular, the marked gain in FL recall suggests a more complete recovery of the dissected lumen, while the simultaneous increase in precision indicates that this improvement was not achieved at the expense of excessive false-positive segmentation. Generalizability on the D4a ASST Lecco clinical cohort (preoperative) was further assessed using masked metrics to account for heterogeneous clinical annotations. The model achieved a masked Dice score of 0.786 for the TL and 0.723 for the FL, with a global masked Dice score of 0.874. Qualitative comparison between predicted segmentations and specialist ground truths confirmed the framework’s ability to resolve the complex topology of the intimal flap even in the presence of variable contrast opacification. Beyond segmentation accuracy, the fully automated pipeline processed a complete case in less than 1 minute on a standard PC workstation (Intel i7-12850HX 16-core CPU, 32 GB RAM, 8 GB GPU), corresponding to an approximately 85% reduction in analysis time compared with manual expert segmentation.

**Table 3:** Comparative analysis on the Graz dataset using out-of-fold predictions. Performance metrics are reported as means. *P* -values were derived from the paired Wilcoxon signed-rank test (significance level 5%) comparing the out-offold AORTA-SCAN predictions against the baseline. Asterisks indicate statistically significant differences.

| **Metric** | **Class** | **nnU-Net** | **AORTA-SCAN** | **Gain (Δ)** | ***p*-value** |
|---|---|---|---|---|---|
| Dice | True lumen | 0.754 | 0.816 | +0.062 | 3.01 × 10<sup>−5</sup>(∗) |
|  | False lumen | 0.707 | 0.837 | +0.130 | 5.26 × 10<sup>−8</sup>(∗) |
| IoU | True lumen | 0.626 | 0.708 | +0.082 | 2.21 × 10<sup>−5</sup>(∗) |
|  | False lumen | 0.590 | 0.748 | +0.158 | 5.26 × 10<sup>−8</sup>(∗) |
| Precision | True lumen | 0.810 | 0.853 | +0.043 | 0.009 (∗) |
|  | False lumen | 0.722 | 0.819 | +0.097 | 2.09 × 10<sup>−7</sup>(∗) |
| Recall | True lumen | 0.732 | 0.796 | +0.064 | 0.005 (∗) |
|  | False lumen | 0.720 | 0.875 | +0.155 | 1.94 × 10<sup>−7</sup>(∗) |

Linear regression analysis (Figure 4, top) demonstrated strong agreement between predicted and reference volumetric measurements. The FL exhibited excellent consistency (Pearson *r* = 0.92), while the TL showed a moderate-to-strong correlation (*r* = 0.79). Residual variability was primarily localized to anatomically complex distal regions, where severe luminal compression, variable contrast opacification, and slit-like morphologies made delineation intrinsically challenging and likely contributed to the remaining discrepancies in TL quantification. The Bland–Altman analysis (Figure 4, bottom) confirmed the absence of substantial systematic bias, with mean differences centered near zero across volumetric scales. Specifically, on the clinical cohort (D4a), the framework maintained stable agreement, with mean RVE values of *−*17.43% for the TL and *−*12.6% for the FL. Among the evaluated biomarkers, the PDI demonstrated the highest clinical stability. AORTA-SCAN achieved a mean absolute PDI error of 0.039, improving upon the 0.087 obtained with the baseline 3D U-Net (*p* = 0.025). Furthermore, the negligible mean PDI bias of *−*0.0017 relative to expert references confirmed the framework’s robustness in preserving volumetric relationships between luminal compartments. By providing stable volumetric measurements that are robust to heterogeneous clinical annotations, the proposed framework established a reliable quantitative baseline for longitudinal TBAD surveillance. Regarding the preoperative ASST Lecco data, the average of the zonal relative PDI biases across the thoracic, visceral, and aorto-iliac regions remained below 17%. Region-specific analysis showed lower variability in the thoracic and visceral regions, while the aorto-iliac segment showed the highest discrepancy (Table 4).

**Figure 4:**
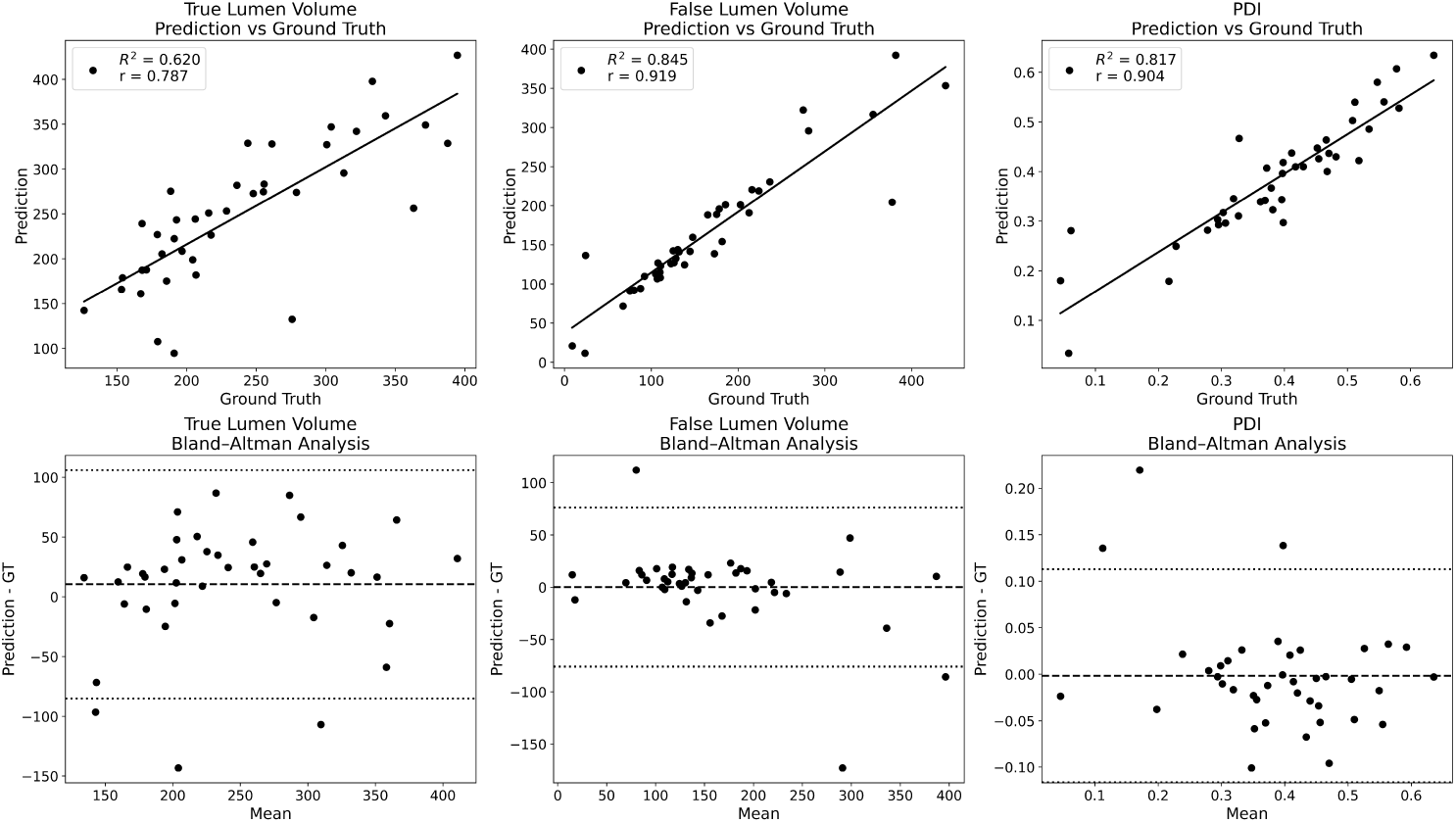
Scatter plots and Bland-Altman diagrams comparing AORTA-SCAN predictions vs expert ground-truth for the Graz University dataset ([22]).

**Table 4:** Global and zonal PDI agreement in the native ASST Lecco dataset. Evaluation was restricted to voxels common to both the ground-truth and predicted lumen masks; therefore, total-volume metrics were not assessed.

| **Region** | **GT PDI** | **Pred. PDI** | **PDI bias** | **PDI MAE** | **PDI SD** |
|---|---|---|---|---|---|
| Global | 0.500 | 0.555 | 11.1% | 0.094 | 0.163 |
| Thoracic | 0.558 | 0.647 | 16.0% | 0.105 | 0.203 |
| Visceral | 0.565 | 0.645 | 14.2% | 0.114 | 0.071 |
| Aorto-iliac | 0.239 | 0.194 | -18.9% | 0.113 | 0.195 |

### 4.2. Postoperative Cohort (ASST Lecco Dataset)

#### 4.2.1. Segmentation Results

Performance on the stented postoperative cohort (*N* = 20) demonstrated the resilience of the triple-input architecture. By isolating metallic components via the high-intensity channel (*>* 600 HU), the framework successfully segmented the stent as a distinct class, achieving a Dice score of 0.723. While this score reflects the inherent sparsity of the metallic mesh, the framework maintained high accuracy for the vascular compartments, achieving a TL Dice score of 0.900 and a FL Dice score of 0.751. This multi-class approach enabled the automated partitioning of the intra-stent compartment, where the TL resides, and the extra-stent space constituted by the residual FL. This helped mitigate beam-hardening artifacts that typically obscure the intimal flap in standard architectures. The clinical reliability of this partitioning was confirmed by a strong Pearson correlation of *r* = 0.96 between predicted and expert TL volumes, supporting the accuracy of the model despite metallic blooming (Fig. 5).

**Figure 5:**
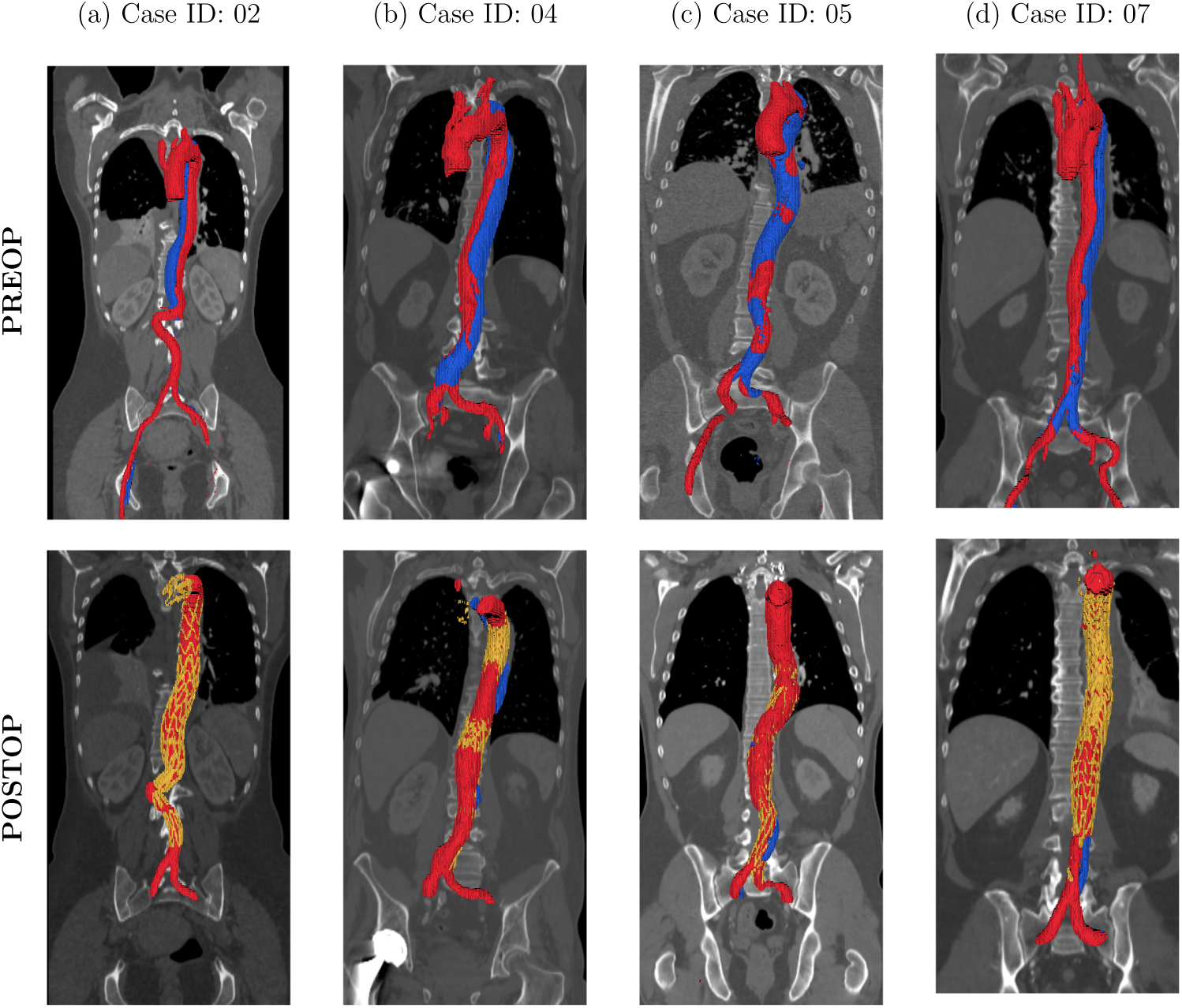
Representative examples of longitudinal TBAD volumetric surveillance. The first row shows four preoperative CTA cases, while the second row shows the corresponding postoperative post-TEVAR examinations from the same patients. The examples illustrate the capability of AORTA-SCAN to preserve anatomically consistent TL/FL segmentation and volumetric characterization across heterogeneous native and postoperative CTA domains.

#### 4.2.2. Clinical Agreement and Volumetric Reliability

The PDI was highly stable in the postoperative setting, with a global mean absolute error of 0.0134. In addition to the global volumetric assessment, a spatially resolved zonal PDI analysis was performed by dividing the aortic volume into three centerline-ordered anatomical regions: thoracic, visceral, and aorto-iliac, as performed by [11]. Across the postoperative cohort, global PDI showed strong agreement between automated predictions and expert references, with a mean ground truth PDI of 0.1177 and a mean predicted PDI of 0.1188, corresponding to a relative PDI bias of 11.39% and a mean absolute error of 0.0134 (Table 5). Zonal analysis showed similarly low errors in the thoracic and visceral regions, with mean absolute PDI errors of 0.0076 and 0.0078, respectively. The aorto-iliac region showed higher variability, with a mean absolute PDI error of 0.0485, consistent with the greater anatomical complexity of distal residual dissection and postoperative remodeling. Despite this regional variability, the global PDI remained stable, supporting its robustness as a volumetric biomarker while suggesting that zonal PDI may provide complementary information for identifying localized discrepancies in FL burden. In the subset of patients with paired preoperative and postoperative CTA scans (*n* = 9), predicted PDI values were compared longitudinally to quantify volumetric remodeling after treatment. The change in PDI was defined as ΔPDI = PDI_preop_ PDI_postop_, so that positive values indicate a postoperative reduction in relative FL burden. Global PDI decreased in all paired cases, with ΔPDI ranging from 0.231 to 0.606 (Table 6). Regionally, the largest reductions were generally observed in the thoracic segment, consistent with the expected effect of proximal endovascular exclusion. In contrast, the aorto-iliac region showed more heterogeneous remodeling, including one case with increased postoperative regional PDI. The framework automatically identified the longitudinal extent of the aorta and extracted equivalent diameter (*D_eq_*) profiles from the global aortic mask, defined as the union of TL and FL segmentations (Table 7). Equivalent diameters were computed on axial cross-sections orthogonal to the extracted aortic centerline, while slices with cross-sectional areas below 50 mm^2^ were excluded to avoid unstable measurements caused by small disconnected components and segmentation noise. The maximum ground-truth *D_eq_* was 39.83 *±* 6.20 mm, compared with 41.49 *±* 6.90 mm obtained with AORTA-SCAN, corresponding to a mean signed error of 1.66*±*3.71 mm and a mean absolute error of 2.56*±*3.15 mm. More spatially robust longitudinal descriptors demonstrated substantially stronger agreement. In particular, the 95*^th^* percentile *D_eq_* exhibited a mean absolute error of only 1.05 *±* 1.67 mm, while the mean *D_eq_* showed the lowest variability, with a mean absolute error of 0.72 *±* 0.66 mm. These findings suggest that percentile-based and longitudinal diameter descriptors are less sensitive to isolated local segmentation irregularities than single-slice maximal measurements (Table 7).

**Table 5:** Aggregated global and zonal PDI agreement in the postoperative ASST Lecco dataset. Values are reported as means across cases. PDI bias is reported as a relative percentage error, PDI MAE is reported in absolute PDI units, and total volume bias is computed as the prediction minus the ground truth (GT).

| **Region** | **GT PDI** | **Pred. PDI** | **PDI bias** | **PDI MAE** | **PDI SD** | **Vol. bias (mL)** | **Vol. MAE (mL)** |
|---|---|---|---|---|---|---|---|
| Global | 0.118 | 0.119 | 11.39% | 0.013 | 0.027 | 0.61 | 15.17 |
| Thoracic | 0.099 | 0.097 | 3.03% | 0.007 | 0.017 | 0.59 | 8.44 |
| Visceral | 0.118 | 0.120 | 1.69% | 0.008 | 0.014 | -0.13 | 1.46 |
| Aorto-iliac | 0.155 | 0.174 | 12.9% | 0.048 | 0.102 | 0.15 | 5.99 |

**Table 6:**
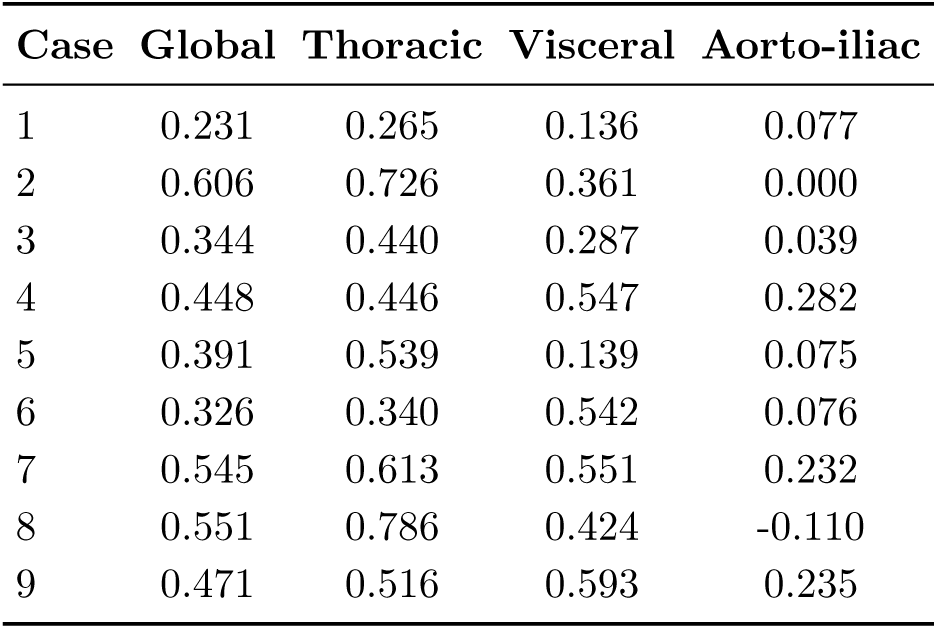
Case-wise longitudinal change in PDI between preoperative and postoperative predicted segmentations computed in 9 patients. Values indicate ΔPDI = PDI_preop_ PDI_postop_, where positive values indicate postoperative reduction in relative FL burden and negative values indicate increased postoperative regional PDI.

**Table 7:** Postoperative equivalent diameter analysis in 20 arterial-phase CTA scans. Values are reported in millimeters as mean *±* standard deviation. *D_eq_* was computed from the global aortic mask, defined as the union of TL and FL labels.

| **Diameter metric** | **Ground Truth** | **AORTA-SCAN** | **Signed error** | **Absolute error** |
|---|---|---|---|---|
| Maximum *D<sub>eq</sub>* | 39.83 ± 6.20 | 41.49 ± 6.90 | 1.66 ± 3.71 | 2.56 ± 3.15 |
| 95th percentile *D<sub>eq</sub>* | 32.49 ± 4.31 | 32.69 ± 3.83 | 0.19 ± 1.96 | 1.05 ± 1.67 |
| Mean *D<sub>eq</sub>* | 24.28 ± 2.80 | 24.57 ± 2.66 | 0.30 ± 0.93 | 0.72 ± 0.66 |
| Standard deviation of *D<sub>eq</sub>* | 6.40 ± 1.18 | 6.17 ± 0.90 | −0.22 ± 0.93 | 0.72 ± 0.63 |

## 5. Discussion

### 5.1. Main findings

The proposed hierarchical framework achieved robust and clinically reliable performance across both native and postoperative TBAD datasets.

In the preoperative setting, the pipeline consistently improved TL and FL segmentation accuracy on both the Graz and ASST Lecco cohorts relative to the baseline architecture, while the postoperative stent-aware configuration preserved high segmentation performance despite severe metallic artifacts and altered vascular anatomy (see Table 3). Volumetric biomarker analysis demonstrated strong agreement with references computed on original surfaces, with the global PDI error remaining below 15% in the Graz cohort and below 12% in both the preoperative and postoperative ASST Lecco cohorts (see Tables 4 and 5). Importantly, the limited global PDI bias confirmed the framework’s ability to preserve the relative volumetric relationship between the true and false lumina, supporting reliable regional and whole-aorta volumetric surveillance. Longitudinal analysis further demonstrated consistent postoperative reduction of the global PDI across paired examinations, reflecting favorable remodeling after treatment, while regional analysis revealed heterogeneous distal remodeling patterns that may provide complementary prognostic information (see Table 6). In addition, equivalent diameter analysis demonstrated good agreement with expert measurements, particularly for longitudinal and percentile-based descriptors, which showed lower variability and greater robustness than single-slice maximal diameter estimation (see Table 7). The complete automated workflow, including segmentation and volumetric analysis, was completed in less than 1 minute on a standard PC-based workstation equipped with an 8 GB GPU.

### 5.2. Hierarchical Strategy and Robustness

The adoption of a hierarchical approach was motivated by the clinical need to decouple global anatomical localization from fine-grained pathological analysis. By leveraging an initial model (SEG1) trained on the entire aorta, the framework isolated the aortic volume from surrounding anatomical structures, such as the spine or kidneys, thus avoiding interference with the following TL and FL segmentation performed by the second-stage models SEG3 (pre-op) and SEG4 (postop). This spatial gating reduced the com-putational burden, allowing the model to process the region of interest at high resolution without the aggressive downsampling often required by fullvolume approaches. The constrained search space enabled the second-stage model to focus its representational capacity on the subtle textural differences between the intimal flap and the two lumina. Such a strategy was particularly relevant in TBAD, where the false lumen may extend over long vascular segments and exhibit heterogeneous contrast enhancement, compression, or partial thrombosis.

### 5.3. Technical and Clinical Significance of Stent Segmentation

The ability to explicitly account for stented anatomy distinguishes the framework from conventional segmentation approaches that are not specifically designed to handle postoperative CTA. The explicit modeling of the stent is clinically relevant because it provides a topological reference for distinguishing the intra-stent compartment from the residual extra-stent space. In postoperative CTA, metallic blooming and beam-hardening can obscure the intimal flap and reduce the reliability of conventional intensity-based segmentation. By incorporating a metal-sensitive representation and treating the stent as an anatomical class, the framework constrains the interpretation of postoperative anatomy and supports more consistent lumen partitioning. This design is particularly relevant for residual TBAD surveillance after TEVAR, where the clinical question is not only whether the aorta can be segmented, but whether the residual FL can be reliably separated from the treated TL compartment. In this context, stent-aware segmentation provides an anatomical constraint that may improve the reproducibility of postoperative volumetric biomarkers and reduce the risk of overestimating luminal volumes in regions affected by metallic artifacts.

### 5.4. Volumetric Biomarkers and Longitudinal Surveillance

The relationship between voxel-wise overlap and volumetric reliability is clinically important. In TBAD, small local segmentation discrepancies may have a limited impact on global volumetric biomarkers, whereas errors in compressed or distal luminal regions can disproportionately affect overlap metrics. For this reason, biomarker agreement should be interpreted alongside traditional segmentation metrics. The PDI is particularly suited to this role because it summarizes the relative contribution of the FL to the total perfused aortic volume and therefore reflects luminal compression and residual disease burden. The zonal formulation of PDI further extends this concept by enabling spatially resolved assessment of remodeling. A global biomarker can summarize overall disease burden, but it may obscure regional differences between the thoracic, visceral, and aorto-iliac segments. This distinction is especially relevant after TEVAR, where proximal exclusion may promote thoracic remodeling while distal residual dissection can persist through re-entry flow or incomplete FL thrombosis. Zonal PDI may therefore provide complementary information for identifying persistent regional FL burden during follow-up. The higher volumetric and PDI variability observed within the aorto-iliac segment (see Tables 4 and 5) compared to the thoracic and visceral regions can be attributed to distinct anatomical and imaging challenges. Specifically, the tapering lumen diameter and the complex tortuosity near the iliac bifurcation inherently increase boundary ambiguity for the segmentation network. Furthermore, the frequent presence of dense peripheral calcifications in this district often may induce blooming artifacts ([15]), which can artificially alter Hounsfield Unit thresholds and obscure the fine separation between the true and false lumens. The automated extraction of equivalent diameter profiles also supports integration with current clinical practice, which remains largely diameter-driven. However, single-slice maximum diameter measurements can be sensitive to local segmentation irregularities, aortic tortuosity, and slice selection. Longitudinal or percentile-based diameter descriptors may provide a more stable representation of vessel caliber along the aortic axis. Taken together, these considerations support the use of automated volumetric and geometric biomarkers as complementary tools rather than replacements for established diameter-based surveillance.

### 5.5. Comparison with Literature

Previous deep learning studies have demonstrated high segmentation accuracy for TBAD analysis on curated CTA datasets. In ([3]), the authors evaluated a serial multi-task CNN on a single-center cohort of 276 preoperative TBAD CTA scans, reporting Dice scores up to 0.93 for TL segmentation and 0.91 for FL segmentation. A multi-stage framework was validated with anatomical straightening on a multicenter dataset of 120 TBAD patients, achieving Dice scores of 0.95 and 0.89 for TL and FL segmentation, respectively ([7]). More recently, a fully automated nnU-Net-based pipeline was developed for AD identification, segmentation, and Stanford classification on 386 CTA scans, reporting Dice scores of 0.968 for TL and 0.971 for FL segmentation ([28]). In the postoperative setting, ([27]) evaluated a global-local fusion framework on a multicenter cohort of 133 patients with 306 postTEVAR follow-up CTA scans, achieving Dice scores of 0.962 for TL and 0.921 for FL segmentation. Although these studies reported slightly higher voxel-wise overlap metrics, they were generally evaluated on highly curated datasets with homogeneous annotations and limited domain variability. In contrast, AORTA-SCAN was specifically designed to operate across heterogeneous real-world clinical domains, including external preoperative cohorts with partial annotations and postoperative CTA scans affected by severe metallic artifacts and altered vascular anatomy. Therefore, the slightly lower segmentation overlap observed in our experiments likely reflects the substantially greater anatomical and clinical variability of the evaluated cohorts. Importantly, despite this increased complexity, the framework maintained stable volumetric biomarker agreement, with the global PDI error remaining below 15% in the Graz cohort and the global PDI bias remaining below 12% in both the preoperative and postoperative ASST Lecco cohorts, supporting the clinical robustness and translational applicability of the proposed approach. Beyond segmentation, several recent studies emphasized the clinical relevance of volumetric biomarkers for TBAD surveillance. **(author?)** [11] demonstrated on a clinical STABILISE cohort of 42 patients that the perfusion dissection index (PDI) can predict postoperative aortic reintervention, while in ([12]) the authors showed in patients with residual aortic dissection that false-lumen volumetric descriptors may outperform diameterbased measurements in predicting adverse remodeling. Similarly, in ([19]) it was reported on a cohort of 40 post-TEVAR TBAD patients that volumetric measurements more accurately characterize postoperative aortic remodeling compared with conventional diameter analysis alone. However, these studies relied on manual or semi-manual volumetric quantification workflows, which remain difficult to integrate into routine large-scale clinical surveillance. In contrast, AORTA-SCAN provides a fully automated end-to-end framework capable of directly extracting volumetric and geometric biomarkers, including PDI and equivalent diameter profiles, from routine CTA scans in both native and postoperative settings, while maintaining clinically stable agreement with expert-derived measurements.

### 5.6. Limitations and Future Directions

This study has limitations that should guide future development. A primary constraint remains the limited availability of high-fidelity annotated data, particularly for postoperative stented cohorts. From a technical perspective, postoperative segmentation in the aortic arch remains challenging, likely reflecting a labeling bias in the training data, where clinical annotations primarily focused on the descending thoracic aorta. Further developments will also refine the spatial analysis of postoperative remodeling by standardizing thoracic, visceral, and aorto-iliac zonal definitions and validating zonal PDI trends in longitudinal follow-up studies. More broadly, AORTA-SCAN establishes the basis for automated longitudinal imaging surveillance following TBAD treatment. By enabling reproducible extraction of volumetric biomarkers across serial CTA examinations, the framework may support standardized follow-up workflows and facilitate earlier identification of adverse remodeling patterns during postoperative monitoring.

## 6. Conclusions

In this work, we developed and validated AORTA-SCAN, a hierarchical deep learning framework for the fully automated characterization of native and postoperative Type B aortic dissection (TBAD). By decoupling global anatomical localization from fine-grained pathological refinement, the framework enables consistent three-dimensional segmentation of the true and false lumina across heterogeneous CTA domains. The integration of stent-aware modeling further supports postoperative surveillance by explicitly accounting for endovascular devices and reducing the impact of metallic artifacts on lumen partitioning. Beyond segmentation, AORTA-SCAN enables automated extraction of clinically relevant volumetric and geometric biomarkers, including TL and FL volumes, PDI, and equivalent diameter descriptors. By reducing the manual burden of volumetric assessment, AORTA-SCAN may support more scalable integration of three-dimensional biomarkers into routine TBAD follow-up.

## Ethics and consent

Ethics approval and consent to participate: Data, provided in pseudoanonymized form, were acquired under the Ethical committed approval entitled STABILISE REGISTRY n. 529, 2019 (Regione Lombardia ASST Lecco).

## Data availability

Data availability: The postoperative ASST Lecco dataset is publicly available at https://drive.google.com/drive/folders/1pGjqeXYNm_SI0jprhsuw0Wre97ARkgrR? usp=sharing.

## Code availability

## Acknowledgments

N/A

## Author contributions

A.M.: Investigation, Methodology, Data analysis, Writing. M:F.: Conceptualization, Data collection, Writing – reviewing and editing. A.M.: Investigation, Data collection. J.J.: Investigation, Data curation. G.R.: Investigation, Data collection. L.M.: Conceptualization, Writing – reviewing and editing. P.C.: Supervision, Investigation, Conceptualization, reviewing and editing.

## Declaration of generative AI and AI-assisted technologies in the manuscript preparation process

During the preparation of this work the authors used Claude (Anthropic) to assist with drafting and editing text and analysis code. After using this tool, the authors reviewed and edited the content as needed and take full responsibility for the content of the published article.

